# Reduced critical care demand with early CPAP and proning in COVID-19 at Bradford: a single centre cohort

**DOI:** 10.1101/2020.06.05.20123307

**Authors:** T Lawton, K Wilkinson, A Corp, R Javid, L MacNally, M McCooe, E Newton

## Abstract

**Background:** Guidance in COVID-19 respiratory failure has favoured early intubation, with concerns over the use of CPAP. We adopted early CPAP and self-proning, and evaluated the safety and efficacy of this approach.

**Methods:** This retrospective observational study included all patients with a positive COVID-19 PCR, and others with high clinical suspicion. Our protocol advised early CPAP and self-proning for severe cases, aiming to prevent rather than respond to deterioration. CPAP was provided outside critical care by ward staff supported by physiotherapists and an intensive critical care outreach program. Data were analysed descriptively and compared against a large UK cohort (ISARIC).

**Results:** 559 patients admitted before 1/May/20 were included. 376 were discharged alive, and 183 died. 165 patients (29.5%) received CPAP, 40 (7.2%) were admitted to critical care and 28 (5.0%) were ventilated. Hospital mortality was 32.7%, and 50% for critical care. Following CPAP, 62% of patients with S:F or P:F ratios indicating moderate or severe ARDS, who were candidates for escalation, avoided intubation. Figures for critical care admission, intubation and hospital mortality are lower than ISARIC, whilst critical care mortality is similar. Following ISARIC proportions we would have admitted 92 patients to critical care and intubated 55. Using the described protocol, we intubated 28 patients from 40 admissions, and remained within our expanded critical care capacity.

**Conclusion:** Bradford’s protocol produced good results despite our population having high levels of co-morbidity and ethnicities associated with poor outcomes. In particular we avoided overloading critical care capacity. We advocate this approach as both effective and safe.

**Social media summary:** The use of early CPAP and proning in COVID-19 was associated with lower critical care admissions, intubation, and mortality at Bradford compared to a large UK cohort (ISARIC WHO CCP-UK).

## Introduction

In early 2020, COVID-19 emerged as a pandemic disease which could result in a severe acute respiratory syndrome. Early respiratory management guidance was drawn up and subsequently revised by both the World Health Organisation (WHO)^1^ and NHS England (NHSE)^2–5^. This strongly favoured early intubation, with NHSE suggesting preparation for intubation of those with a respiratory rate of ≥20 breaths per minute and oxygen saturations of ≤94% despite treatment. Continuous positive airway pressure (CPAP) treatment was deemed appropriate in only select patients and later as a ceiling of care or a bridge to intubation, rather than an ongoing management strategy.^2,6^

Bradford is a deprived^7^ and ethnically diverse UK city, with 32.5% of the population non-white.^8^ It has high rates of comorbidity, particularly diabetes with the highest prevalence in the UK (10.8% vs UK 6.9%).^9^ All these factors are associated with worse COVID-19 outcomes.^10,11^ Bradford Royal Infirmary serves a population of approximately 500,000, with 16 critical care beds capable of supporting invasive ventilation, normally staffed as 8 ICU/8 HDU. As the COVID-19 crisis unfolded, an additional ICU was opened, expanding this to 28 beds. However, early modelling suggested that 30% of hospital admissions might require invasive ventilation.^12^ Plans were made for a third ICU, but there were concerns about staffing, swamping of hospital infrastructure, and potentially unnecessary early invasive ventilation with its attendant risks to patients and the system.

In mid-March Qin Sun et al. argued that a combination of risk stratification, early critical care admission, CPAP, and awake prone positioning, could result in reduced intubation rates and possibly improve mortality.^13^ Other early publications reported high numbers of patients managed on non-invasive ventilation (NIV).^14–17^ Experiences from Italy and China suggested that high levels of intubation rapidly saturated critical care capacity, leading to worse outcomes and highlighting the need to prevent unnecessary intubation.^18,19^

One concern was critical care staffing, given the complexity of managing an intubated patient in full personal protective equipment (PPE) even assuming a bed was available. Another was the potential limitation of oxygen supplies with high use by some equipment.^20^ We therefore acquired a large number of air-driven CPAP machines (DeVilbiss SleepCube), and set them up as “Fixed CPAP” devices with entrained oxygen for use in the early treatment of severe COVID-19. As simple devices intended for out of hospital use, they were readily acceptable to ward staff with support, whilst bringing some benefits of critical care treatment to the wards.

Concerns regarding the use of NIV have hinged on viral aerosolisation, potential lack of efficacy, and confusion between BIPAP and CPAP regarding harmful overdistension.^1^ Studies of disease transmission with NIV appear largely based on unfiltered exhalation ports.^21–23^ Initial concerns about efficacy reflect findings in MERS, in which a study reported 5 NIV failures.^24^ However, it is unclear whether CPAP or BIPAP was used - and neither was a denominator provided.

There are reasons that CPAP may benefit COVID-19 patients who do not require immediate intubation. Unlike most causes of ARDS, lungs affected by COVID-19 can remain compliant and recruitable in early illness, with work of breathing remaining low in comparison to hypoxia caused by atelectatic changes.^25^ CPAP can provide a sustained positive airway pressure, without increasing tidal volume and remaining lung protective.^26^

Prone positioning of intubated patients with severe ARDS now forms part of standard recommendations.^27^ Early awake proning with NIV has also been found to be beneficial, leading to reduced intubation rates.^28^ More recently, self proning has improved oxygen saturations in COVID-19 patients in emergency care.^29^

Bradford therefore adopted the widespread early use of CPAP and self proning in the management of more severe COVID-19, with the aims of improving patient outcomes and controlling critical care demand. It was not feasible to admit all patients requiring CPAP to the critical care unit and an ‘ICU without walls’ approach became necessary. Our initial experience and outcomes are presented below.

## Methods

This single centre retrospective cohort study was conducted at Bradford Royal Infirmary, a teaching hospital in the UK.

### Intervention

Our approach was designed and delivered by a multidisciplinary team comprising doctors from critical care and respiratory, acute and emergency medicine, together with nursing staff and physiotherapists. It comprised elements including awake proning, escalation planning, and usual critical care therapies. However, the core intervention was the use of early CPAP in hypoxaemic respiratory failure due to COVID-19. This required a massive expansion of our capacity to deliver CPAP outside critical care.

As well as 21 existing NIV machines, 100 “Fixed CPAP” machines were used with entrained oxygen. HME viral filters were added prior to the expiratory port, and they were used with non-vented masks. We considered helmet use but were unable to obtain any due to supplies being restricted to Italy. The “Fixed CPAP” machines were introduced on 3rd April 2020 in anticipation of a peak in demand a week later. This expansion ensured that all patients who might benefit from CPAP could be offered it, even if they were not considered appropriate for critical care escalation.

A dedicated critical care outreach consultant was available 24 hours a day, undertaking twice daily outreach ward rounds. Patients on CPAP received daily respiratory physiotherapy sessions, and physiotherapists oversaw most of the CPAP provision. Nurses and physiotherapists with experience in NIV were seconded to CPAP wards, providing additional support.

The protocol for managing respiratory failure in patients with confirmed or suspected COVID-19 is outlined in Figure 1, and was initiated as soon as possible either in ED, AMU or one of two designated CPAP inpatient wards. Transfers on CPAP were rare, but did occur with cleared corridors. Despite the use of HME viral filters, the use of CPAP was considered to be aerosol generating and occurred only in designated ‘red zones’ which were mostly open wards and wards with small bays, where staff used standard ‘airborne’ PPE including FFP3 mask. Nasal high-flow oxygen was used on occasion for CPAP rest periods. Patients not receiving CPAP were treated with low flow oxygen therapy devices, typically nasal cannula or humidified venturi devices.

**Figure 1.**
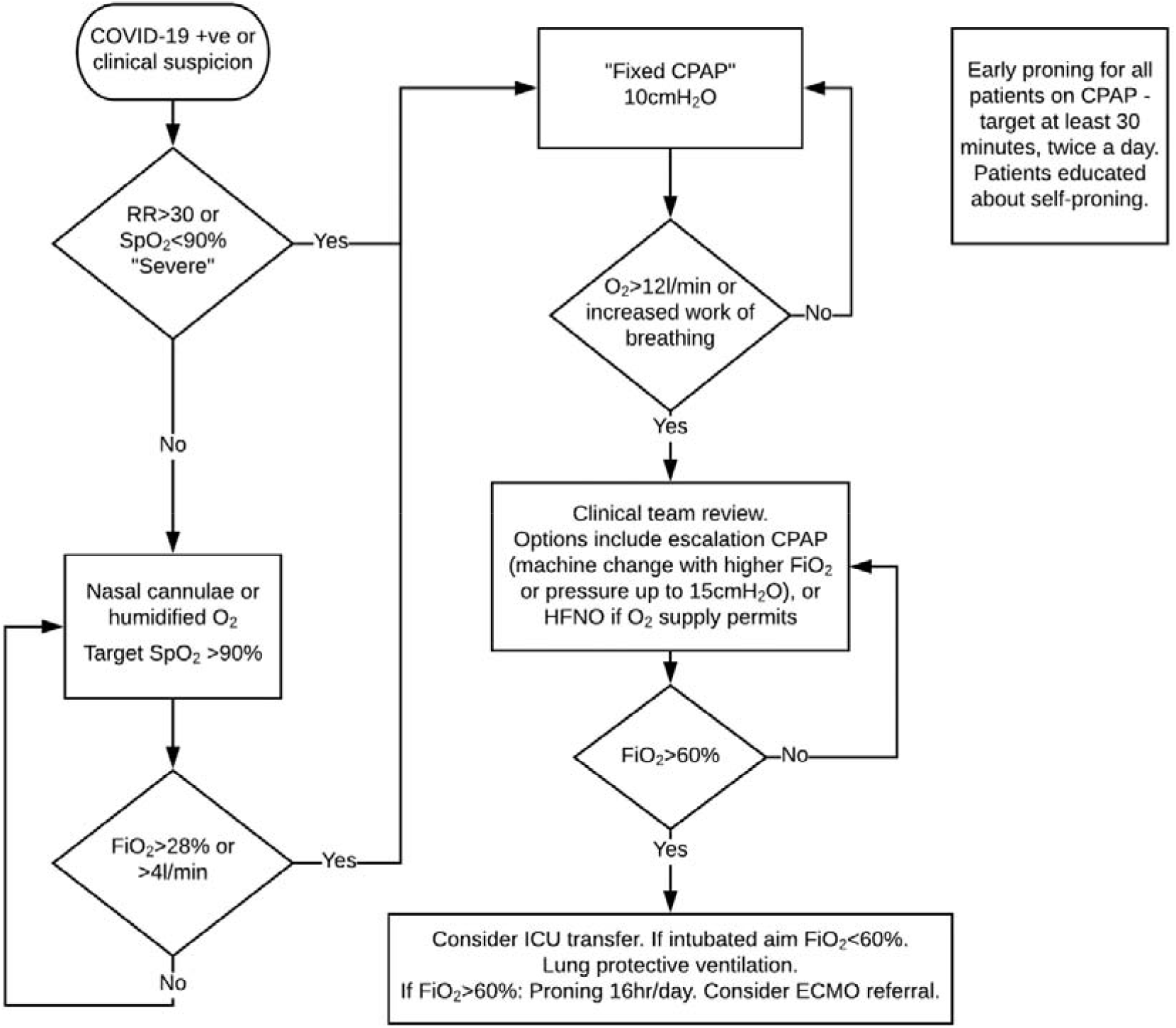
COVID-19 respiratory protocol.

Patients were educated about the benefits and indications for proning if able to comprehend and physically able to self-prone. Patients receiving CPAP were encouraged to prone for at least 30 minutes twice a day - in practice usually for a few hours.

Early discussion and documentation of decisions on critical care escalation by the admitting team was encouraged in line with GMC guidance. This decision was made on an individual patient basis and was not age-based or protocolised. Compliance with this was excellent, aided by a COVID-19 proforma in the electronic patient record. Escalation to critical care for patients deemed appropriate for intubation involved continuation of CPAP, in an environment prepared for rapid intubation should it be required.

During the study period, Bradford was a recruiting site for the RECOVERY trial investigating treatments for COVID-19.

### Data

We included all patients with a positive COVID-19 PCR test admitted prior to 1st May 2020, and other patients where the treating team considered COVID-19 the most probable diagnosis – any repeat testing was the decision of the admitting team. Patients receiving CPAP were identified separately by daily review of wards capable of delivering NIV. The first patient was diagnosed on 26^th^ February 2020 and data was collected from all patients to discharge or death. As an audit of practice reporting data only in aggregate, the need for formal ethical approval and consent was waived by our research department.

A retrospective review of the Electronic Patient Record (Cerner Millennium) was conducted. Demographic, admission and outcome data were collected for all patients. Selected co-morbidities, ceilings of care, critical care admissions and escalation to invasive ventilation were also recorded. Observations were recorded at first presentation of COVID-19 (respiratory rate, pulse oximetry, arterial gases, inspired oxygen therapy).

Where CPAP was used, the initial machine was documented together with duration and any escalation of therapy. Observations before and after initiation of initial and escalation CPAP were recorded, as well as at the point of maximum support. Where CPAP was not used, maximum oxygen therapy was recorded. Partially because many patients were proning themselves, proning was often not recorded specifically in the notes. This limited analysis of this component, but self-proning remained part of the treatment protocol during the entire period.

For audit purposes only, patients were retrospectively assessed against two sets of intubation criteria based on widely publicised advice for COVID-19. Firstly, a respiratory rate ≥20 combined with oxygen saturations ≤94% and ≥15L/min oxygen or equivalent, as advocated by NHS England on 26th March 2020^3^ and still current.^4^ Secondly, a ratio of arterial oxygen partial pressure to FiO_2_ (P:F) of <200mmHg (26.6kPa), as recommended explicitly in German guidelines,^30^ and implicitly in other guidelines recommending NIV is only used in mild ARDS.^31^ These assessments were made prior to the use of CPAP (or at admission in patients who did not receive it), and at the point of highest respiratory support during CPAP use. As arterial lines were only used in patients admitted to critical care, arterial gas results were not available for all patients. Where they were unavailable, a P:F ratio of <200mmHg was taken to be equivalent to an oxygen saturation to FiO_2_ (S:F) ratio of <214%.^32^ We attempted to avoid false positives on this criterion by excluding patients with oxygen saturations above 94%, and using the lowest equivalent S:F ratio we found in reliable literature.^33^ The FiO_2_ of variable performance devices was also calculated conservatively, resulting in estimated oxygen concentrations lower than cited in the literature.^34^ During the period described, we did not use formal criteria for intubation decisions, which were made by the critical care team in conjunction with the relevant medical teams.

Analysis was largely descriptive. Comparison was made with the ISARIC WHO CCP-UK cohort of 20133 patients admitted to UK hospitals,^35^ noting that this cohort also contains data from Bradford. Outcomes were compared using the 2-tailed exact binomial test and the sign test for medians. Data were analysed in R 4.0.0 (R Core Team, 2020).

## Results

559 patients were included in the cohort, of whom 165 received CPAP (29.5%). 376 were discharged from hospital alive and 183 died, one of whom had been transferred for ECMO. All patients are included in the analysis. A flow diagram for the cohort is given in Figure 2. Demographics and comorbidity results for the cohort and subgroups are given in Table 1. The cohort was mostly male (54.7%) with a median age of 68. Rates of obesity (35.4%) and diabetes (33.6%) were high, and as risk factors for severity they were even higher in the groups treated with CPAP and invasive ventilation. The proportion of all measured comorbidities in the cohort exceeded the ISARIC average, reflecting the overall poorer state of Bradford’s health.

**Table 1.** Demographics and comorbidities.

|  | **Bradford CPAP (n=165)** | **Bradford Intubated (n=28)** | **Bradford patients (n=559)** | **ISARIC patients<sup>35</sup>** |
| --- | --- | --- | --- | --- |
| Sex Male | 58.2% | 75% | 54.7% | 59.9% |
| Median Age (IQR) (range) | 64 (53-72) (18-92) | 57 (49-64) (31-70) | 68 (53-81) (0-102) | 72 (58-82) (0-104) |
| Hypertension | 51.8% | 46% | 51.3% | 45.3%<sup>36</sup> |
| Cardiac conditions | 35.8% | 31% | 38.6% | 30.9% |
| Obesity | 42.7% | 48% | 35.4% | 10.5% |
| Respiratory conditions | 42.1% | 21% | 34.2% | 32.1% |
| Diabetes | 40.6% | 39% | 33.6% | 28.1% |
| CKD | 27.6% | 11% | 31.0% | 16.2% |
| Smoking | 15.8% | 21% | 11.5% | 6.0% |
| Cancer | 7.9% | 11% | 10.8% | 10.4% |

Comorbidity data is near complete except obesity (84% complete)

**Figure 2.**
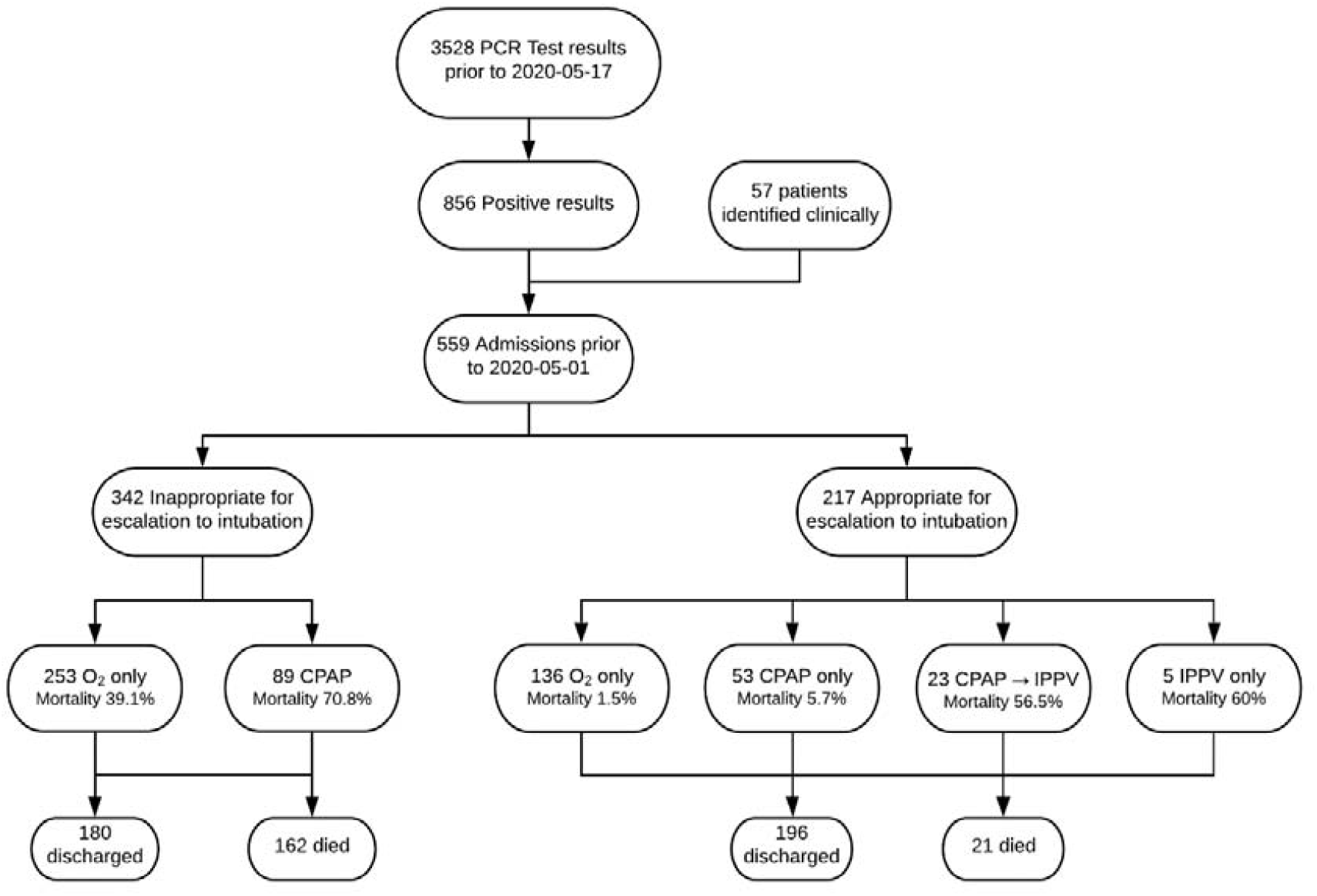
Study flow chart.

Respiratory parameters for the cohort receiving CPAP are given in Table 2, with values immediately before and after starting initial CPAP treatment. Over half of these patients (51.5%, 85 of 165) had evidence of at least moderate ARDS by P:F or S:F criteria before starting treatment. Changes given are for the group; on a post-hoc examination of individual data there did appear to be a division between “responders” and “non-responders” to CPAP. Whole cohort outcomes are given in Table 3. Bradford had a markedly lower critical care admission (7.2%) and intubation (5.0%) rate than the ISARIC cohort, with comparable hospital mortality overall (32.7%) and for critical care patients (50%). Estimating from the ISARIC values, Bradford would have expected 55 patients requiring intubation and 92 critical care admissions. We intubated 28 patients from 40 admitted to critical care. Of these, 23 had been treated with NIV prior to intubation. We had a peak occupancy of 16 COVID-19 patients in critical care for a total of 21 patients. The third ICU was not required. A small number of patients initially considered appropriate for intubation died without being intubated. This was largely due to changes in escalation plans, or in two cases sudden cardiac arrest.

**Table 2.** Respiratory parameters, CPAP group (n=165) – median (IQR)

|  | Before CPAP | After starting CPAP |
| --- | --- | --- |
| SpO<sub>2</sub> | **92%** (90-96) | **94%** (92-96) |
| FiO<sub>2</sub> | **47%** (32-60) | **50%** (37-60) |
| Respiratory rate | **28** (24-34) | **28** (23-32) |
| **Patients with arterial gases available (n=39)** |  |  |
| SpO<sub>2</sub> | **92%** (90-94) | **94%** (92-96) |
| FiO<sub>2</sub> | **60%** (44-60) | **60%** (45-75) |
| Respiratory rate | **32** (26-40) | **28** (23-34) |
| P:F ratio | **14.9kPa** (12.1-20.3) | **18.6kPa** (12.4-25.0) |
| PaO<sub>2</sub> | **7.6kPa** (7.0-8.8) | **8.8kPa** (7.9-11.1) |

**Table 3.** Outcomes.

|  | Bradford patients | ISARIC patients | p |
| --- | --- | --- | --- |
| NIV use | **29.5%** (165 of 559) | **15.9%** (2670 of 16805) | <0.0001 |
| Critical care admission | **7.2%** (40 of 559) | **16.5%** (3001 of 18183) | <0.0001 |
| Intubation | **5.0%** (28 of 559) | **9.8%** (1658 of 16866) | <0.0001 |
| Hospital mortality | **32.7%** (183 of 559) | **38.6%** (5165 of 13364) | 0.005 |
| Mortality for critical care patients | **50%** (20 of 40) | **53.7%** (958 of 1784) | NS |
| Median Length of Stay (IQR) | **7** (3-14) | **7** (unreported IQR)<sup>36</sup> | NS |

Results for the assessment of all patients against the two described intubation criteria are detailed in Tables 4 and 5. In the group appropriate for intubation, the majority of patients (78.9%, 60 of 76) receiving CPAP had S:F or P:F evidence of moderate or severe ARDS at some point during their stay and would have required intubation on some guidelines. However most of these patients (61.7%, 37 of 60) were treated with only CPAP and avoided intubation. Patients who met these criteria but avoided intubation had a relatively low mortality (8.1%, 3 of 37). In the group where intubation was not appropriate, meeting intubation criteria was associated with high mortality.

**Table 4.** Intubation criteria - group appropriate for intubation.

|  | Timing | Met criteria | Intubated at any point | Overall Mortality | Mortality where not intubated |
| --- | --- | --- | --- | --- | --- |
| *SpO<sub>2</sub> ≤ 94% & RR ≥ 20 & FiO<sub>2</sub> ≥ 60%* | Prior to any CPAP | 7.8% (17 of 217) | 64.7% (11 of 17) | 35.3% (6 of 17) | 16.7% (1 of 6) |
|  | Worst during CPAP | 42.1% (32 of 76) | 62.5% (20 of 32) | 43.8% (14 of 32) | 25.0% (3 of 12) |
| *P:F ratio <200mmHg or equivalent* | Prior to any CPAP | 19.4% (42 of 217) | 38.1% (16 of 42) | 26.2% (11 of 42) | 7.7% (2 of 26) |
|  | Worst during CPAP | 78.9% (60 of 76) | 38.3% (23 of 60) | 26.7% (16 of 60) | 8.1% (3 of 37) |

**Table 5.** Intubation criteria - group not appropriate for intubation.

|  | Timing | Met criteria | Hospital Mortality |
| --- | --- | --- | --- |
| *SpO<sub>2</sub> ≤ 94% & RR ≥ 20 & FiO<sub>2</sub> ≥ 60%* | Prior to any CPAP | 12.3% (42 of 342) | 83.3% (35 of 42) |
|  | Worst during CPAP | 62.9% (56 of 89) | 80.4% (45 of 56) |
| *P:F ratio <200mmHg or equivalent* | Prior to any CPAP | 19.3% (66 of 342) | 84.8% (56 of 66) |
|  | Worst during CPAP | 83.1% (74 of 89) | 75.7% (56 of 74) |

During preparation of this paper, ISARIC WHO CCP-UK released updated data which has been used in Tables 1 and 2.^35^ It is worth noting that the earlier ISARIC data^36^ had a lower proportion of NIV use (12.1%) and higher critical care admission rate (18.6%), which may reflect changing practice in the UK as opinion on CPAP use developed.

## Discussion

This cohort demonstrates that this approach can avoid the need for invasive ventilation in many patients who would have been intubated on official guidelines. However, this is an uncontrolled cohort study, and evidence from randomised controlled trials will be required to determine whether CPAP is best instituted early, or used as a rescue therapy in the case of deterioration. Whilst we used early CPAP and proning at Bradford, it is plausible our results could have been achieved by using CPAP only later on. However this would have probably necessitated more critical care admissions even if intubation were avoided. Response to CPAP must be rapidly and repeatedly assessed as some patients will still require invasive ventilation, and patients with COVID-19 can deteriorate quickly.

It is difficult to unpick the contribution of the main treatments offered as part of our protocol: early CPAP and self-proning, particularly as we lack data on the exact timing of self-proning. However self-proning is a relatively easy and uncontroversial addition to early CPAP and we saw many examples of rapid improvement as a patient lay on their front; we regard our results as due to the protocol in its entirety.

Our approach involved turning wards into “red zones” with full airborne PPE including FFP3 masks; additionally filters were used on CPAP expiration ports. We do not have data on staff sickness rates by area, but anecdotally during this period it was lower on wards where airborne PPE was used.

Whilst the majority of patients were identified from lists of positive PCR results, we also included patients being treated as COVID-19 on clinical grounds. Our close supervision of patients receiving CPAP mean we may have been more likely to identify these patients than if they were elsewhere in the hospital. We tried to avoid this by accessing data from the hospital command centre, though this depended on teams reporting COVID-19 suspicions centrally.

We chose to compare against ISARIC as the largest UK hospital dataset with detailed comorbidity and outcome data. As a product of research-active hospitals we expect its results to be as good as or better than average.^37^ Bradford’s data being included in the ISARIC cohort will tend to dilute the differences seen.

Assessment against the two sets of intubation criteria showed a large proportion of patients who fulfilled criteria but were treated with CPAP instead. A number of patients were quickly put onto CPAP as a first therapy and improved, thus never fulfilling the criteria relating to oxygen use. Also, the point of highest support recorded in the data was not always where the P:F ratio was lowest. We therefore regard the numbers reported as an underestimate of those who might have fulfilled criteria had our CPAP protocol not been in use. The high rates of meeting those criteria in the group treated with CPAP reflect that our protocol was selecting patients with more severe COVID-19 for CPAP treatment.

Other centres have reported similar results in smaller cohorts of patients treated with CPAP. Of 24 patients in Liverpool appropriate for intubation, 38% were intubated,^38^ similar to our figure of 30% of 76 patients. In Newcastle, of 28 patients who were not appropriate for escalation there was a 50% mortality^39^ – our higher figure of 71% from 89 patients may represent differing patient selection or random variation. A multicentre report from Italy of 157 patients reported a 44% failure rate (defined as intubation or death);^40^ our failure rate was 54% overall – 71% and 34% in the non-escalation and escalation groups respectively. This may relate to their exclusion criteria removing some of the more severely unwell patients; our goal was to offer CPAP to all who might possibly benefit.

Despite a greater burden of comorbidities than the ISARIC cohort, and serving a population expected to have poor outcomes, Bradford managed a much lower critical care admission and intubation rate with a comparable or lower mortality. Many patients who would have required invasive ventilation under early guidance were able to recover on CPAP without the exposure to the multiple potential harms resulting from invasive ventilation on the critical care unit. Intubated patients would be expected to stay longer, decondition more, and suffer more iatrogenic lung injury so CPAP may have reduced morbidity in the longer term. Equally, the comparable mortality rate for patients admitted to critical care suggests that the use of early CPAP to prevent intubation did not result in harm where it only delayed deterioration.

## Conclusions

This approach is relatively low cost and low tech. By reducing ventilator demand it does not rely on a surplus of highly trained staff, nor on a generous oxygen supply. As such we consider it may have wider applicability outside the UK healthcare system.

At the time of submission, we consider the first wave to be concluded in Bradford. Our second ICU is currently closed. Based on our experience, we intend to continue early CPAP during any second wave, and we would recommend other centres consider the use of CPAP and proning in any patient with more severe COVID-19.

## Data Availability

The datasets generated during and/or analysed during the current study are available from the corresponding author on reasonable request.

## Acknowledgements

We thank Kirstin McGregor, Peter Szedlak, Margaret Aslet, Rosa Gallie, Matthew Bromley, Laura Stephenson, Daniel Cummings, and Caroline Bonner (all BTHFT) for their work on data collection; and the BTHFT physiotherapy, respiratory, A&E, critical care, and acute medicine teams for their work with protocols and supporting this project.

## Research in context

### Evidence before this study

We searched PubMed and medRxiv for articles published between January 2020 and the start of this study delineating the use of early continuous positive airway pressure (CPAP) in the treatment of COVID-19, using the search terms (“covid” or “covid-19” or “coronavirus”) and (“CPAP” or “NIV” or “prone” or “proning”). We found several case series documenting the use of NIV, but only one paper describing the principles of systematic early CPAP and proning leading to reduced rates of mechanical ventilation. However, this study contained little detail on the delivery of CPAP therapy and also described a low threshold for ICU admission. We found no published accounts of widespread CPAP use outside critical care.

### Added value of this study

To our knowledge, this is the largest observational study to date to feature an in-depth exposition of early CPAP and proning outside critical care. 559 COVID-19 patients were included, with 165 receiving CPAP. Our analysis demonstrates favourable rates of critical care admission, intubation and mortality. Many patients who met previously recommended intubation criteria were successfully managed without this, moreover reported outcomes were no worse where intubation was ostensibly delayed for a trial of non invasive ventilation. Additionally, we furnish a detailed account of our pragmatic and multi-disciplinary approach which we hope may be of interest to fellow clinicians, either as a model to manage further waves of COVID-19, or alternatively to free up critical care capacity for resumption of pre-COVID hospital activity.

### Implications of all the available evidence

This dataset adds considerably to a growing body of evidence that early CPAP and proning can safely be recommended as a treatment strategy for COVID-19, reducing exposure to the risks of sedation and mechanical ventilation. Its widespread delivery can be organised in a resource-efficient manner to avoid overwhelming hospital capacity.

### STROBE Checklist

STROBE Statement—Checklist of items that should be included in reports of ***cohort studies***

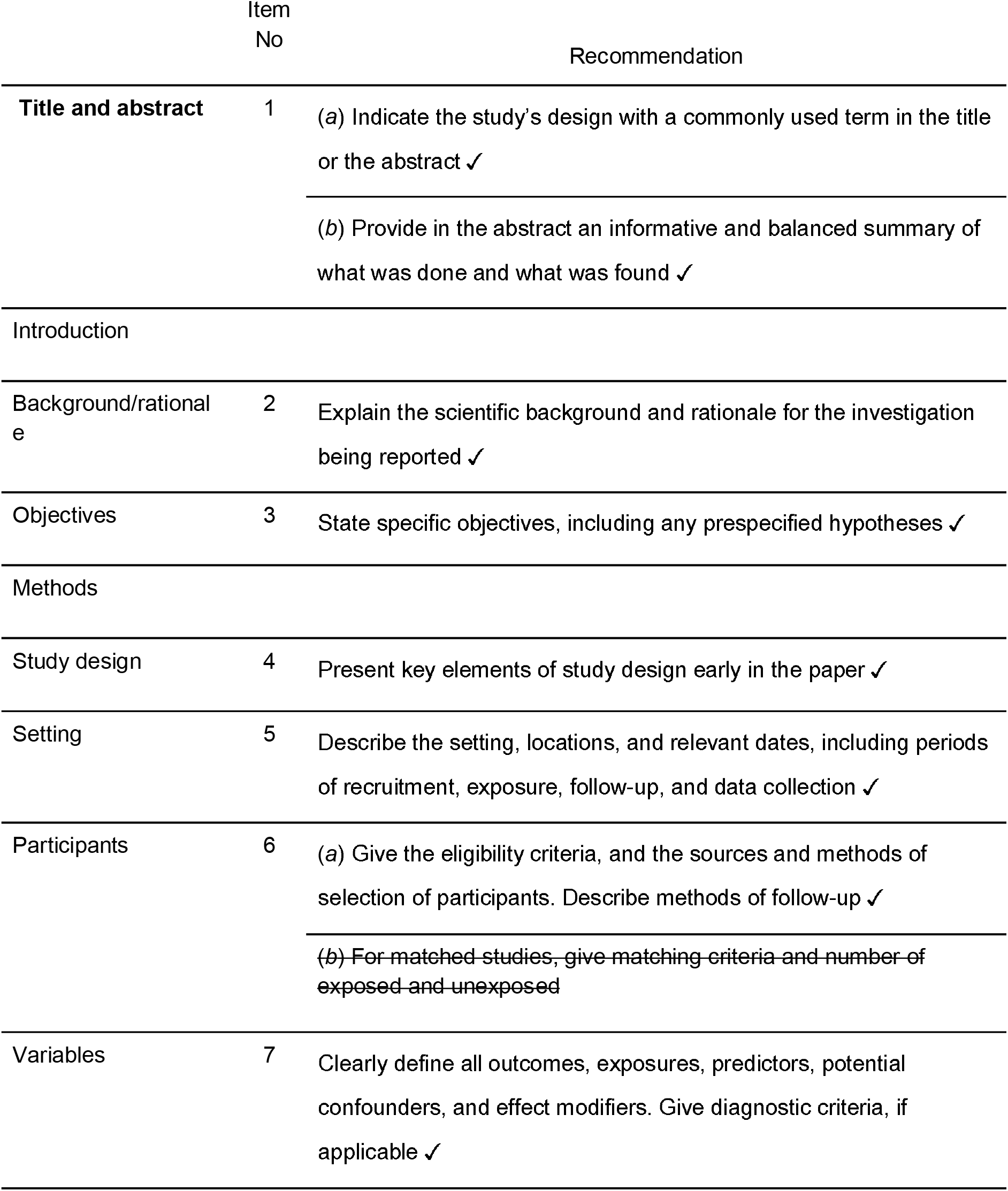
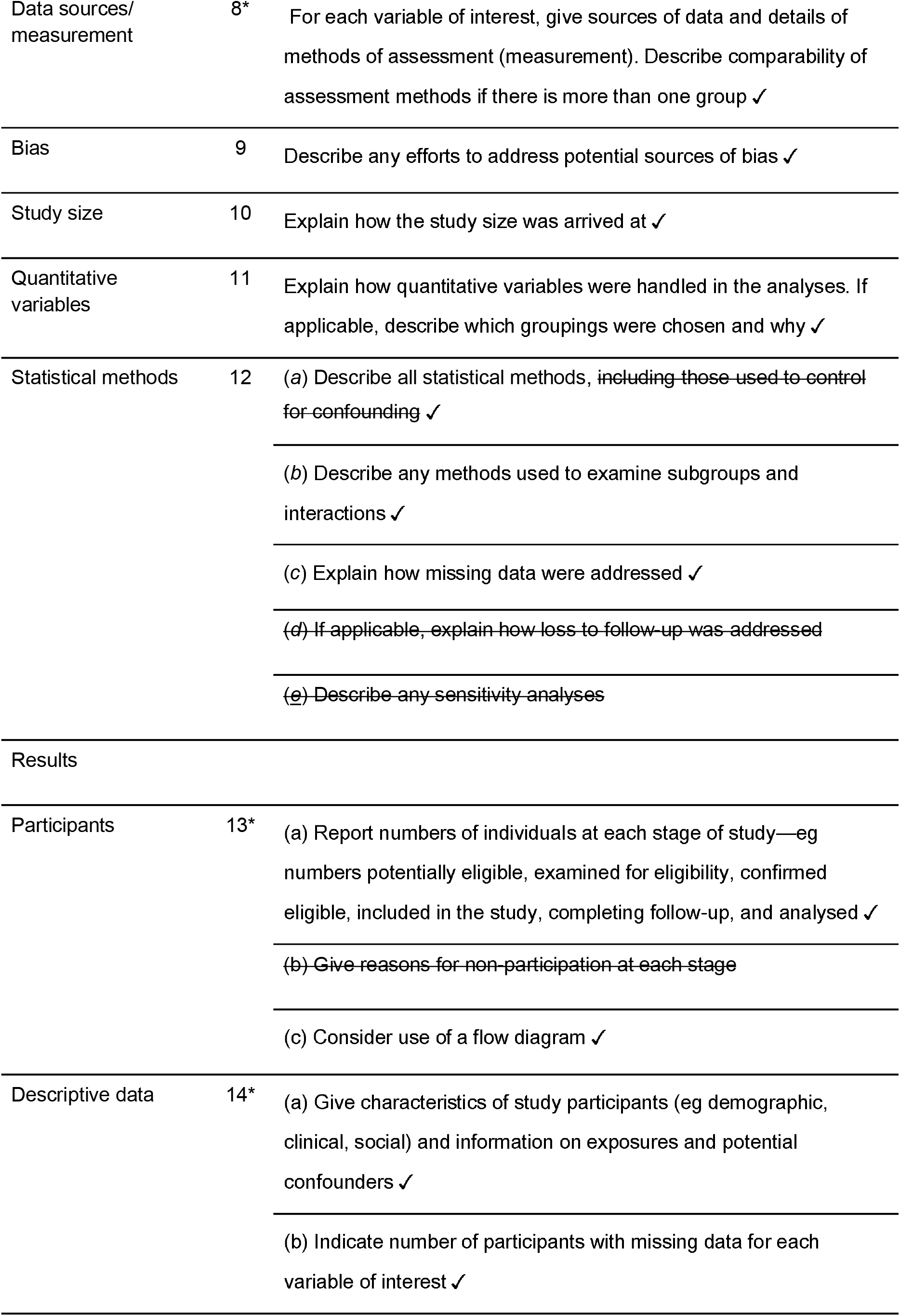
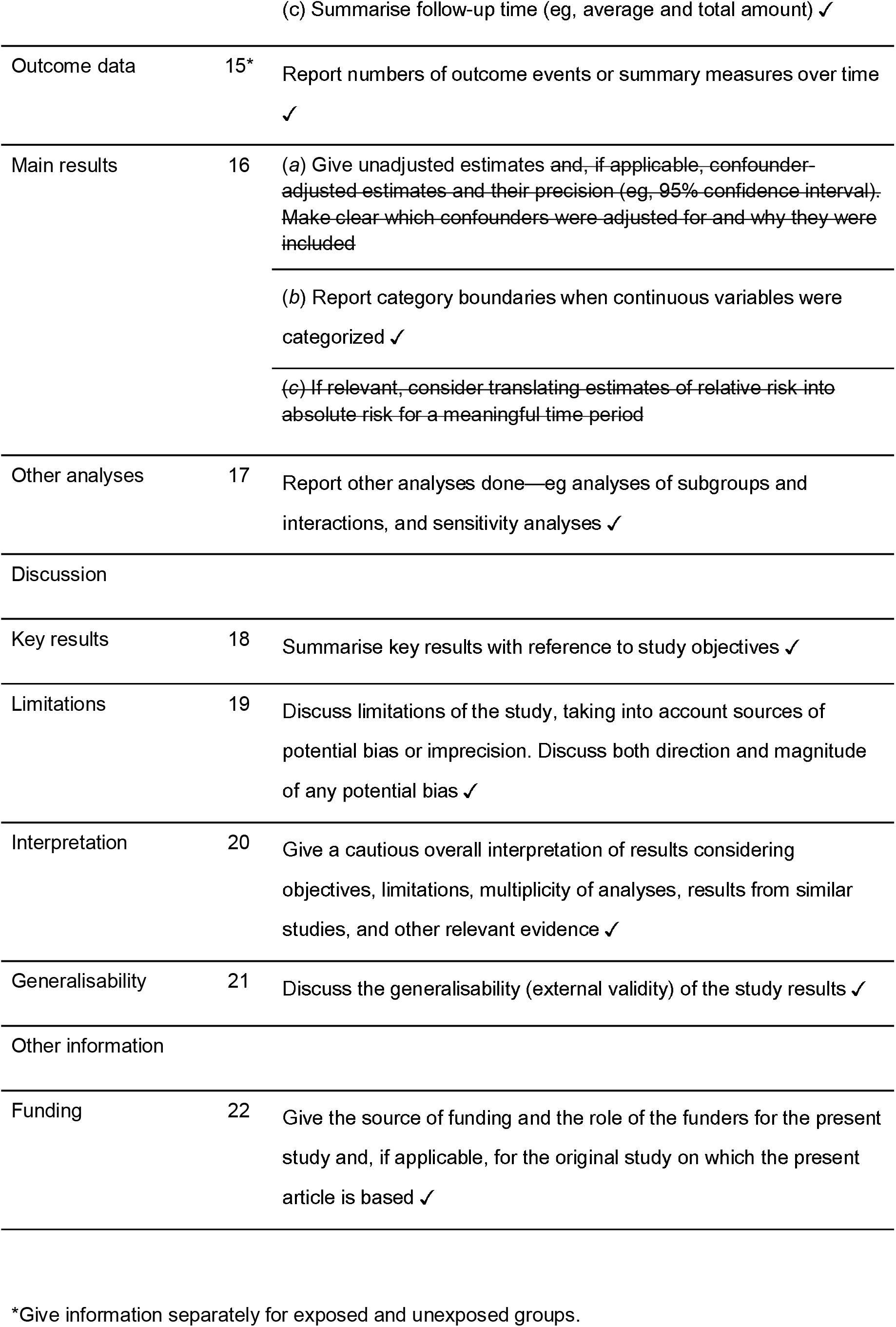
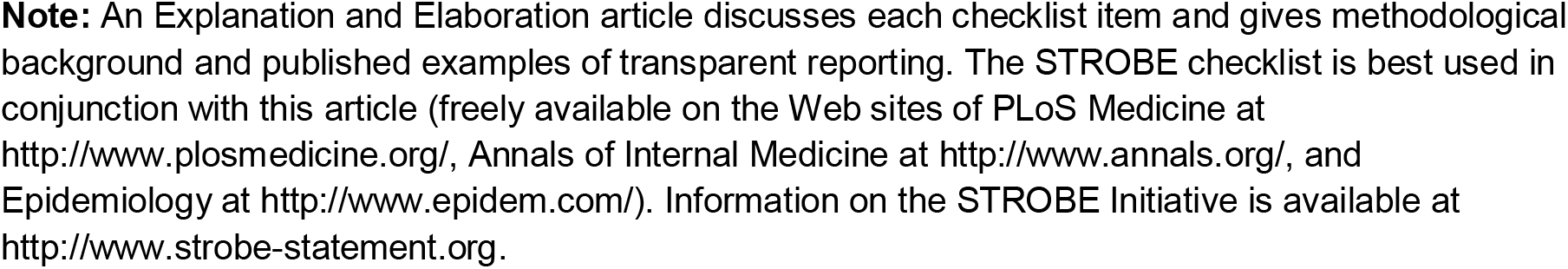

